# Isolation of *Borrelia puertoricensis* from ticks collected in Merida, Yucatan, Mexico

**DOI:** 10.1101/2024.07.22.24310794

**Authors:** Edwin Vázquez-Guerrero, Alexander R. Kneubehl, Guadalupe C. Reyes-Solís, Carlos Machain-Williams, Aparna Krishnavajhala, Paulina Estrada de los Santos, Job E. Lopez, J. Antonio Ibarra

**Affiliations:** Departamento de Microbiología, Escuela Nacional de Ciencias Biológicas, Instituto Politécnico Nacional, Ciudad de México, México; Department of Pediatrics, Baylor College of Medicine, Houston, Texas, USA; Centro de Investigaciones Regionales Dr. Hideyo Noguchi, Universidad Autónoma de Yucatán, Mérida, México; Department of Molecular Virology and Microbiology, Baylor College of Medicine, Houston, Texas, USA

## Abstract

Isolation of tick-borne relapsing fever (TBRF) spirochetes has proven to be a useful tool to understand their distribution by geographic areas where the tick vectors inhabit. However, their isolation and culture are not easy and in general an animal model is needed to achieve this task. Here, argasid ticks were collected from a neighborhood in Ciudad Caucel, Merida, Mexico, and *Borrelia puertoricensis* was isolated with an immunosuppressed mouse model. By using this model, a higher number of spirochetes were observed in blood samples, and these were successfully cultivated in Barbour-Stoenner-Kelly (BSK)-IIB media. Genomic analyses confirmed that the isolate to be *B. puertoricensis*. The present report shows that this spirochete is present in argasid ticks in Ciudad Caucel and presents a potential medical and veterinary health risk.

**Author Summary:** Relapsing fever is a neglected disease caused by spirochetes bacteria of the *Borrelia* and genus that are different than those in the Lyme disease group. These bacteria are transmitted mostly by *Ornithodoros* (*Alectorobious*) soft ticks. In many countries, including Mexico, *Ornithodoros* species are understudied and often misidentified. In this report we obtained an isolate of *Borrelia puertoricensis* from ticks captured in a small park in the city of Merida, Yucatan. These results emphasize the need for fine-scale vector surveillance in neighborhoods to determine the abundance of *Borrelia* species, and to define their impact on humans and domestic and wild animals.

## Introduction

Relapsing fever (RF) is caused by pathogenic spirochetes in the genus *Borrelia* and are primarily vectored in the Western Hemisphere by argasid ticks. These vectors feed quickly, within five to 60 minutes, and then return to the nest, den, or under floor tiles and windowsills inside houses [1,2]. Argasid ticks have a life cycle that complicates the detection of the *Borrelia* species they transmit. Specially, the genus *Ornithodoros* feed within five to 60 minutes and subsequently return to the cavity in which they dwell. This includes wild animal nests or dens [1]. In the American continent TBRF is caused by *Borrelia hermsii*, *Borrelia parkeri*, *Borrelia turicatae, Borrelia mazzottii,* and *Borrelia venezuelensis*, which are all transmitted by different species of argasid ticks, while *Borrelia miyamotoi* is transmitted by ixodid ticks [3]. However, in Latin America, the distribution of argasids and the RF spirochete species they transmit has been unclear.

While RF spirochetes were described in Mexico in the twentieth century, the disease is not recognized by healthcare providers [4]. Humans infected with TBRF spirochetes show nonspecific symptoms that include sporadic fever, headache, myalgias, chills, nausea, vomiting, rigors, and pregnancy complications [5]. TBRF is often misdiagnosed as Lyme disease, although no isolates of the latter pathogen have been described in Latin America.

There has been taxonomic confusion with argasid ticks from Latin America, causing a poor understanding of the public health impact of RF borreliosis. For example, in the early 1900s, a species of RF spirochete circulated in Panama causing significant morbidity [6]. *Ornithodoros talaje* was implicated as the vector [7], but recent records of this tick from Panama are absent. Another argasid from Panama that was not considered as the vector of RF spirochetes was *Alectorobius* (*Ornithodoros*) *puertoricensis,* which has previously been misidentified as *O. talaje*. A comparative analysis of *O. talaje* and *A. puertoricesis* by Fox indicated that the two were synonymous [8]. Venzal’s work further showed that adults and nymphs of *O. talaje* and *A. puertoricensis* are morphologically similar [9]. Recent collections of *A. puertoricensis* from Panama demonstrated that this species transmits *Borrelia puertoricensis* [2]. Collectively, these studies indicated that updated surveillance efforts are needed for *A. puertoricensis*.

In Mexico, *A. puertoricensis* has been described in the state of Veracruz [10], but it is unknown whether *B. puertoricensis* circulates in the country. We report surveillance efforts in the state Yucatan, Mexico, and collected ticks in neighborhoods of Merida. Using an immunosuppressed mouse model, we describe the isolation of a *Borrelia* species from *A. puertoricensis* ticks. The genomic characterization of this spirochete identified it as *B. puertoricensis*. We also performed a comparative analysis of the diagnostic antigen *Borrelia* immunogenic protein A (BipA) across known species of RF spirochetes [11]. Our results demonstrated that *B. puertoricensis* extends to the southeastern part of Mexico and are found in areas with high human activity.

## Methods

### Sampling sites

In January 2023, argasid ticks were collected from a small park in the Ciudad Caucel neighborhood in Merida city (latitude 20.985954, longitude -89.695441), the capital of Yucatan (Fig 1A). Opossum (*Didelphis virginiana*) were sighted in the neighborhood and four of their burrows were sampled using an aspirator or dry ice as a source of carbon dioxide to bait ticks, as previously described [12] (Fig 1B, 1C). Collected samples were placed in 50-ml ventilated centrifuge tubes and ticks from each burrow were kept separated.

**Fig 1.**
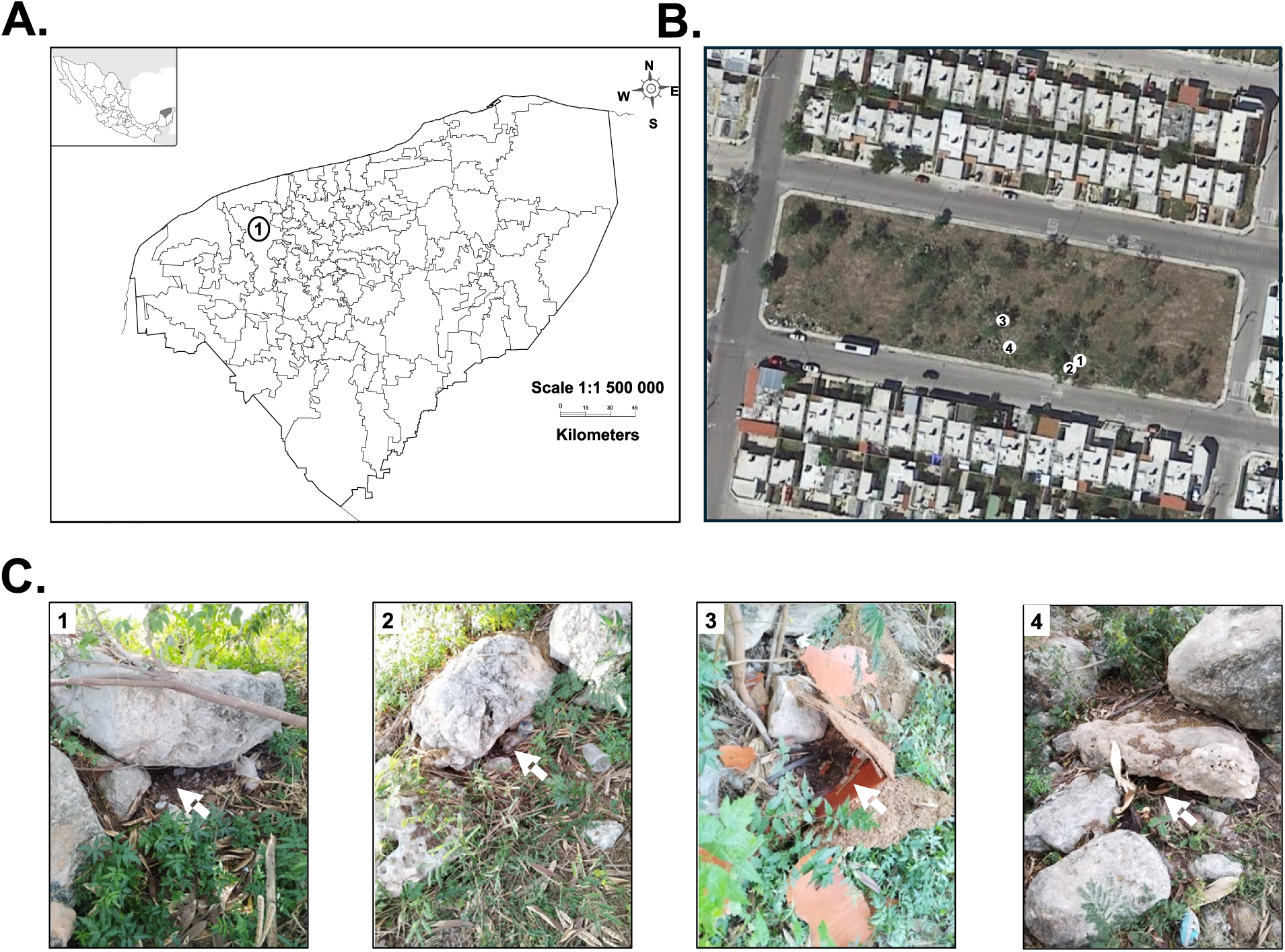
Collection and identification of *Alectorobius* (*Ornithodoros*) *puertoricensis* in Yucatan, Mexico. Shown is a map of the state of Yucatan, the number 1 in the map shows the location of Merida city, left upper corner map shows the location of Yucatan in Mexico (A). Collection site was in Ciudad Caucel, here in a terrain map and the numbers indicate location of the burrows (B). The white arrows in (C), panels 1 through 4, point to where ticks were collected. In (C), C1 to C4 point to the respective locations in panel (B). Terrain maps were obtained from Google maps and used under the “principles of fair use” (https://about.google/brand-resource-center/products-and-services/geo-guidelines/).

### Argasid tick identification and classification

Live ticks were returned to our laboratory in Mexico City and maintained at 25 °C and 85% relative humidity inside a glass desiccator as described previously [11]. Ticks were speciated by using light microscopy and previously described taxonomical guides [13, 14, 15]. Molecular classification was done as follows: genomic DNA was isolated from ticks (n = 3) with a DNeasy Blood and Tissue kit following the manufacturer’s instructions (Qiagen, Hilden, Germany). A ∼475 nucleotide region of the mitochondrial 16S rDNA gene was amplified by PCR as previously described [16, 17] using the Tm 16S+1 (5’-CTGCTCAATGATTTTTTAAATTGC-3’) and Tm 16S-1 (5’-CCGGTCTGAACTCAGATCATGTA-3’) primers. Amplification conditions were performed, as previously described [16, 17]. PCR products were observed in 1% TAE agarose gels and further submitted for sequencing to obtain a 2X coverage at Macrogen Inc. (https://dna.macrogen.com/). Sequences were assembled and trimmed using MUSCLE v5 and ChromasPro (https://technelysium.com.au/wp/chromaspro/) [18]. Consensus sequences obtained were compared with available sequences in GenBank using BLASTn (https://blast.ncbi.nlm.nih.gov). Nucleotide identity was assigned based on the expected value (e-value). In cases of identical e-values, we assigned the gene to the sequence with the highest value. A phylogenetic analysis using the maximum likelihood method was performed in PhyML 3.1 with the GTR+G model [19]. Sequences were deposited in GenBank.

### Animal model

All animal procedures were approved by our Institutional Animal Care and Use Committee (CEI-ENCB protocol # ZOO-001-2022e1) To determine whether the collected ticks were infected by *Borrelia* spp. a mouse model was used as described previously with modifications [12]. Ticks were groups in pools and fed on one of three different mouse strains (DBA/2J, C57BL/c or BALB/c) depending on the availability at the Instituto Politécnico Nacional. In total, 27 animals were used. Two and a half microliters of blood were collected from mouse tails for 18 consecutive days to evaluate the presence of spirochetes by dark field microscopy (Olympus CX33 Microscope), as described previously [2]. Spirochetes were counted per 6 μl of blood (dilution 1:2 with EDTA 1%) under dark-field illumination by using a Petroff–Hausser counting chamber (Hausser Scientific, Horsham, PA) with a depth of 0.02 mm [2]. When spirochetes were observed, the infected mouse was euthanized by exsanguination to quantify and isolate the bacteria as described [2]. If no bacteria were observed the mice were kept alive and after 30 days, the animals were exsanguinated. The blood samples were centrifuged at 5,400 x *g* for two minutes to obtain the sera. Serum samples were used to evaluate antibody responses to *Borrelia* by immunoblotting, as described below.

Given that isolation of bacteria was not possible with the previous protocol even when spirochetes were observed we opted to use a previously chemically induced immunosuppression model [20, 21]. Briefly, seven and four days prior to feeding ticks, 10 mg/kg dexamethasone was subcutaneously administered to mice. Dexamethasone was also administered twice a week during the experiment and the presence of spirochetes was determined as described above. After seven days mice were exsanguinated, blood was processed as described [2], and BSK-IIB media was inoculated with infected serum.

### Serology

Immunoblotting was performed to assess seroconversion of mice to *Borrelia* recombinant glycerophosphodiester phosphodiesterase (rGlpQ). The recombinant protein was purified as described [22], electrophoresed, and transferred to polyvinylidene fluoride (PVDF) membranes (Millipore). Immunoblotting was performed, as previously described [22]. Serum samples were diluted 1:200 and used to probe the PVDF membranes. HRP-conjugate recombinant protein G (diluted 1:4,000) was used to detect IgG. Antibody binding was detected by chemiluminescence using Novex ECL Western Blotting Detection Reagent (Invitrogen) and immunoblots were imaged with a ChemiDoc MP (Bio-Rad, Hercules, California, USA).

### Bacterial culture

For spirochete isolation, when bacteria were observed in murine blood, the animal was exsanguinated by intracardiac puncture and whole blood was centrifuged at 500 x *g* for 5 min. The plasma was removed and centrifuged again at 5,000 x *g* for 10 min [23]. The pellet was resuspended in 1 ml of BSK-IIB media and used to inoculate 4 ml of medium [24], supplemented with 10 µg/ml rifampin, 4 µg/ml phosphomycin, and 0.5 µg/ml amphotericin B [25]. Cultures were incubated at 35°C and 5% CO_2_ for approximately eight days. To assess bacterial growth, an aliquot of the culture was placed on a glass slide and observed by dark field microscopy. When bacteria were detected, they were subcultured in fresh BSK-IIB media for storage and DNA isolation purposes.

### Bacterial identification and genomics

Spirochetes were grown for no more than two passages, and bacterial genomic DNA was isolated using a phenol-chloroform procedure. Pulsed field electrophoreses was performed to determine DNA quality, as described [26]. For an initial identification, a PCR for the gene coding for the 16S rRNA and *flaB* was performed; Polymerase chain reaction (PCR) conditions consisted of initial denaturation at 95°C for 2 min followed by 35 cycles at 95°C for 30 sec, annealing at 55°C for 30 sec, and an extension at 72°C for 2 min; after the last cycle, a final extension was performed at 72°C for 5 min [17]. To confirm the expected molecular size for each amplicon, PCR products were observed in 1% TAE agarose gel electrophoresis and further submitted for sequencing to obtain a 2X coverage at Macrogen Inc. (https://dna.macrogen.com/). Sequences were assembled and trimmed using MUSCLE v5 [18] and ChromasPro (https://technelysium.com.au/wp/chromaspro/). Consensus sequences obtained were compared with available sequences in GenBank using BLASTn (https://blast.ncbi.nlm.nih.gov). Nucleotide identity was assigned based on the expected value (e-value). In cases of identical e-values, we assigned the gene to the sequence with the highest value. A phylogenetic analysis using the maximum likelihood method was performed in PhyML 3.1 with the GTR+G model [18]. Sequences were deposited in GenBank.

To determine plasmid content and DNA integrity 1 μg of purified genomic DNA was subjected to pulse field electrophoresis as previously described [17]. Long-read sequencing was performed using the Oxford Nanopore Technologies (ONT) Mk1B platform with the SQK-RBK114.96 library preparation kit and R10.4.1 flow cell. Short-read sequences were generated by SeqCenter (SeqCenter, Pittsburgh, Pennsylvania, USA) using an Illumina 2x150 library preparation kit. Plasmid-resolved genome assembly was generated by using short-reads to polish the long-read data, as previously reported [27] with modification. Due to the increase in accuracy for Oxford Nanopore Technologies’ platform and to prevent over polishing of the assembly, which can introduce errors, we modified the polishing pipeline for genome assembly. The mean ONT coverage was 439x and the mean Illumina coverage was 236x. Using a previously established approach [27], completeness and QV scored (based on the Phred scale) were determined to be 99.89% and Q53.82, respectively. The assembly was annotated with NCBI’s Prokaryotic Genome Annotation Pipeline and submitted to NCBI’s GenBank (accession numbers CP149102-CP149123).

## Results

### Capture and identification of Alectorobius (Ornithodoros) puertoricensis

Surveillance for argasid ticks was conducted in neighborhood parks in Ciudad Caucel, located in the at the outskirts of west side of the Merida city (Fig 1A), capital city of the Mexican state of Yucatan (892,000 persons by the 2020 national census). Opossum (*Didelphis virginiana*) burrows were in a small park in the center of the neighborhood (Fig 1B) and 466 soft ticks were collected from four burrows (Fig 1C, Table 1). The life stages included 279 nymphs, 28 larvae, and 159 adults (56 females and 103 males) (Table 1).

**Table 1.**
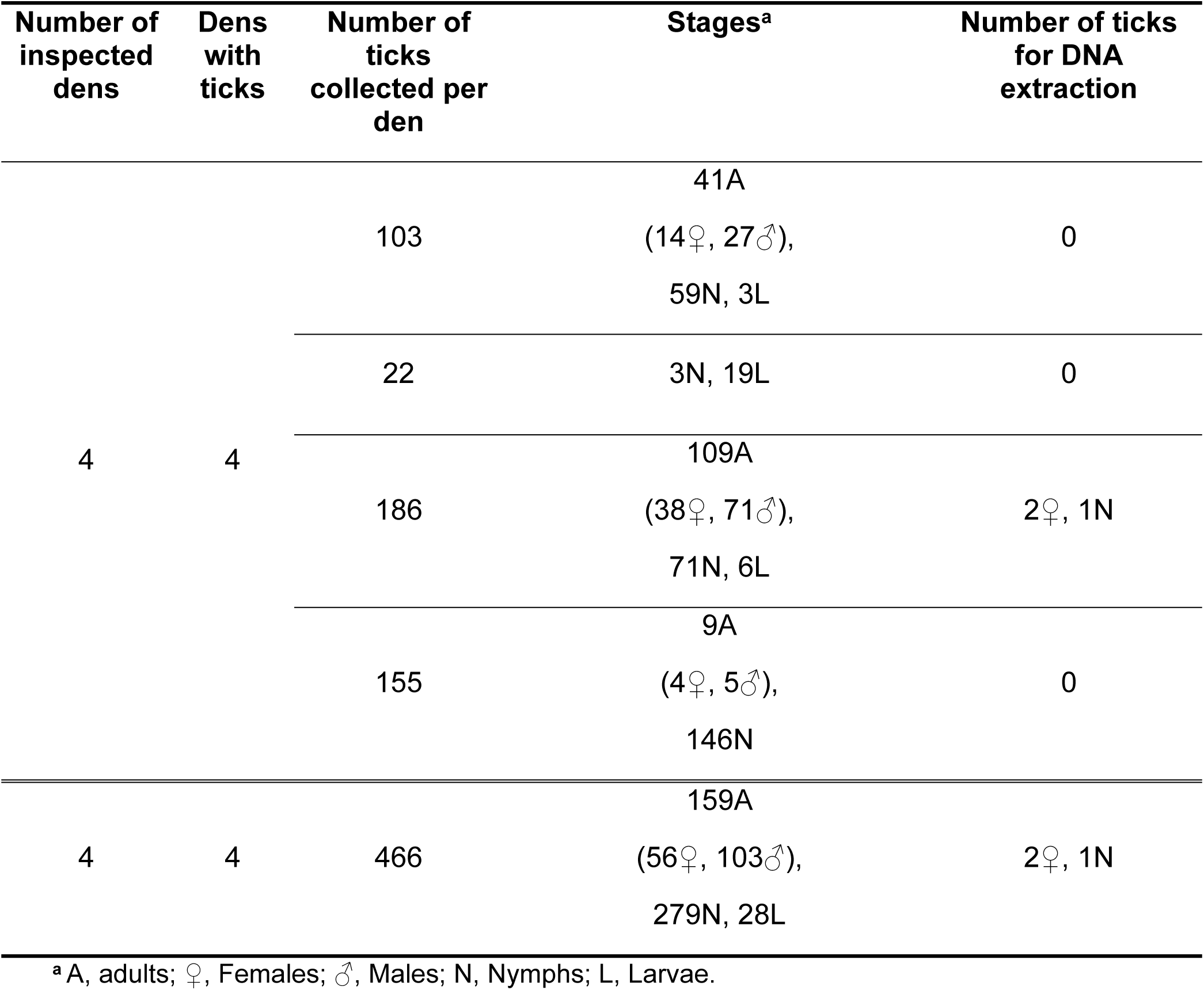
Characteristics of collected ticks.

Morphological identification of adults and nymphs indicated that all collected ticks were *A. puertoricensis*. The morphology was consistent with the original descriptions of Fox (1947) and with specimens previously reported in a Mexico [15]. These included the absence of eyes, cheeks present, body oval, hood present and well developed, and tarsus I with subapical dorsal protuberance (Fig 2A, 2B).

**Fig 2.**
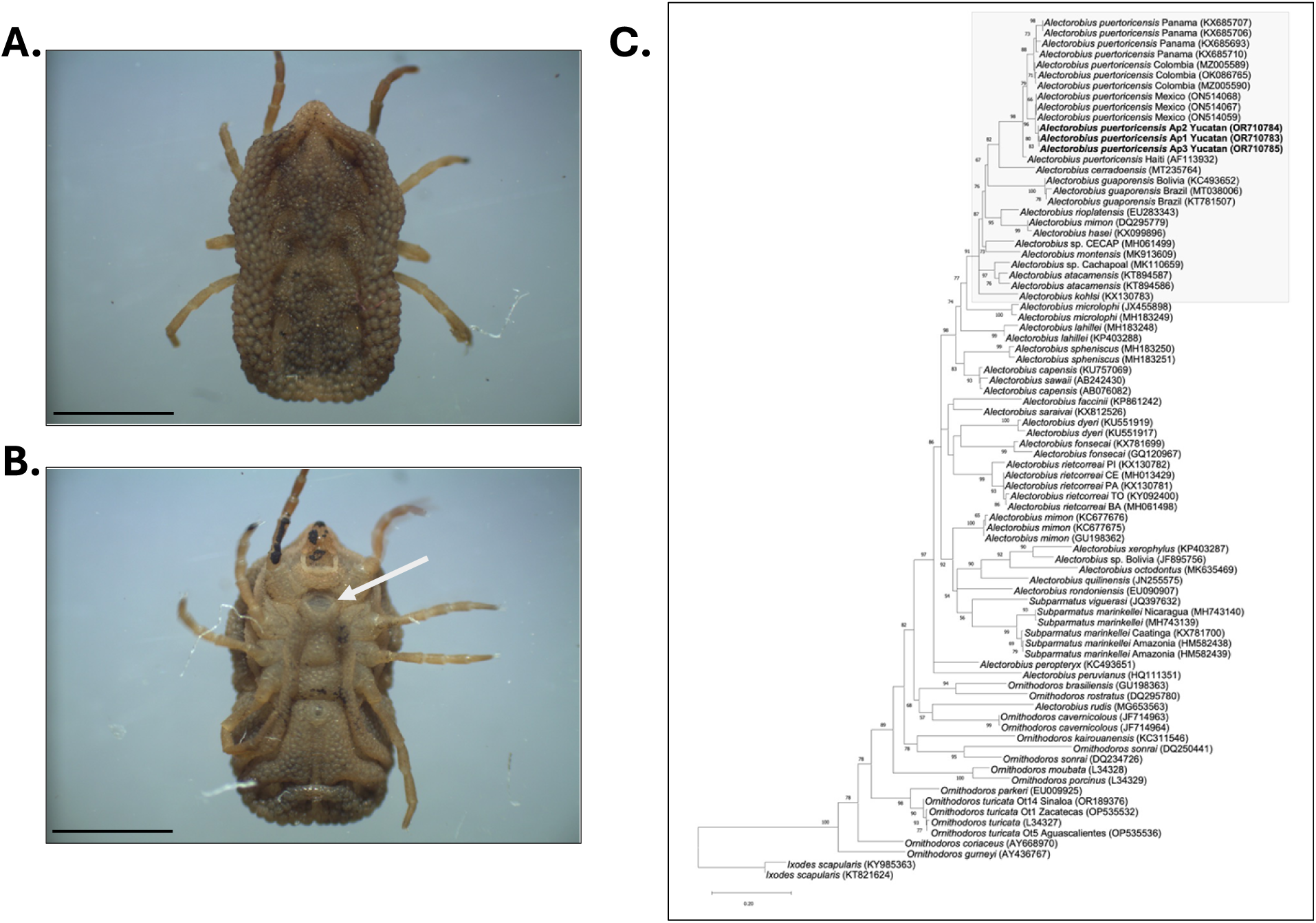
Morphological and molecular identification of *A. puertoricensis*. Stereo microscope images of a female *A. puertoricensis* specimen. The dorsal view is shown in (A) and the ventral in (B). The white arrow points to the genital aperture. Scale bars: 3 mm. In (B), a phylogenetic analysis of mitochondrial DNA sequence fragments from soft ticks *Alectorobius* spp is shown. The analysis was performed with the maximum likelihood method using the GTR+G model. Bootstrap is shown in the branches. Shown in bold letters are the sequences of the analyzed ticks. In the gray box is the group *talaje*. In parenthesis the accession numbers at NCBI. The bar displays the differences between sequences. *Ixodes scapularis* was used as an outgroup.

Morphological speciation was complemented with molecular identification by analyzing 16S rDNA mitochondrial sequences. Total DNA from three ticks was obtained and analyzed (Ap1 to Ap3). BLASTn analysis indicated that sequences from the three samples were ∼97% identical to *A. puertoricensis* from multiple regions of Central America and the Caribbean. The sequences were deposited in GenBank (OR710783-OR710785) and phylogenetic analyses of all three sequences corroborated that they grouped with *A. puertoricensis,* and they are in the same group as those reported from Mexico, Panama, Colombia, and Haiti (Fig 2C). These results show that *A. puertoricensis* is present in Mexico likely parasitizing opossums in urban neighborhoods.

### Isolation of Borrelia puertoricensis

To determine whether the collected ticks were colonized by relapsing fever (RF) spirochetes, we fed them on mice. We observed a low level of spirochetemia (2.5 x 10^4^ bacteria per ml of blood) in four mice and failed to successfully culture the bacteria in Barbour-Stoenner-Kelly (BSK)-IIB medium. The four cohorts of ticks that were infected were subsequently fed on mice under chemical immunosuppression. We used one DBA/2J mouse and three BALB/c mice. In these mice we observed higher bacterial densities of 5 x 10^5^ to 5 x 10^6^ spirochetes per ml of blood. After seven days the four mice were exsanguinated, blood was processed as described [2], and BSK-IIB media was inoculated with infected serum. We successfully grew spirochetes that originated from the DBA/2J mouse in medium and generated glycerol stocks. This isolate was stored at -70 °C and designated CAU1 (*B. puertoricensis* Ciudad <u>Cau</u>cel isolate 1). DNA was extracted from the cultures and used for PCR amplification and sequencing of the *16S* rRNA coding gene and *flaB,* which indicated that the spirochetes were *B. puertoricensis* (Fig 3). Given these findings, we performed a whole genome analysis.

**Fig 3.**
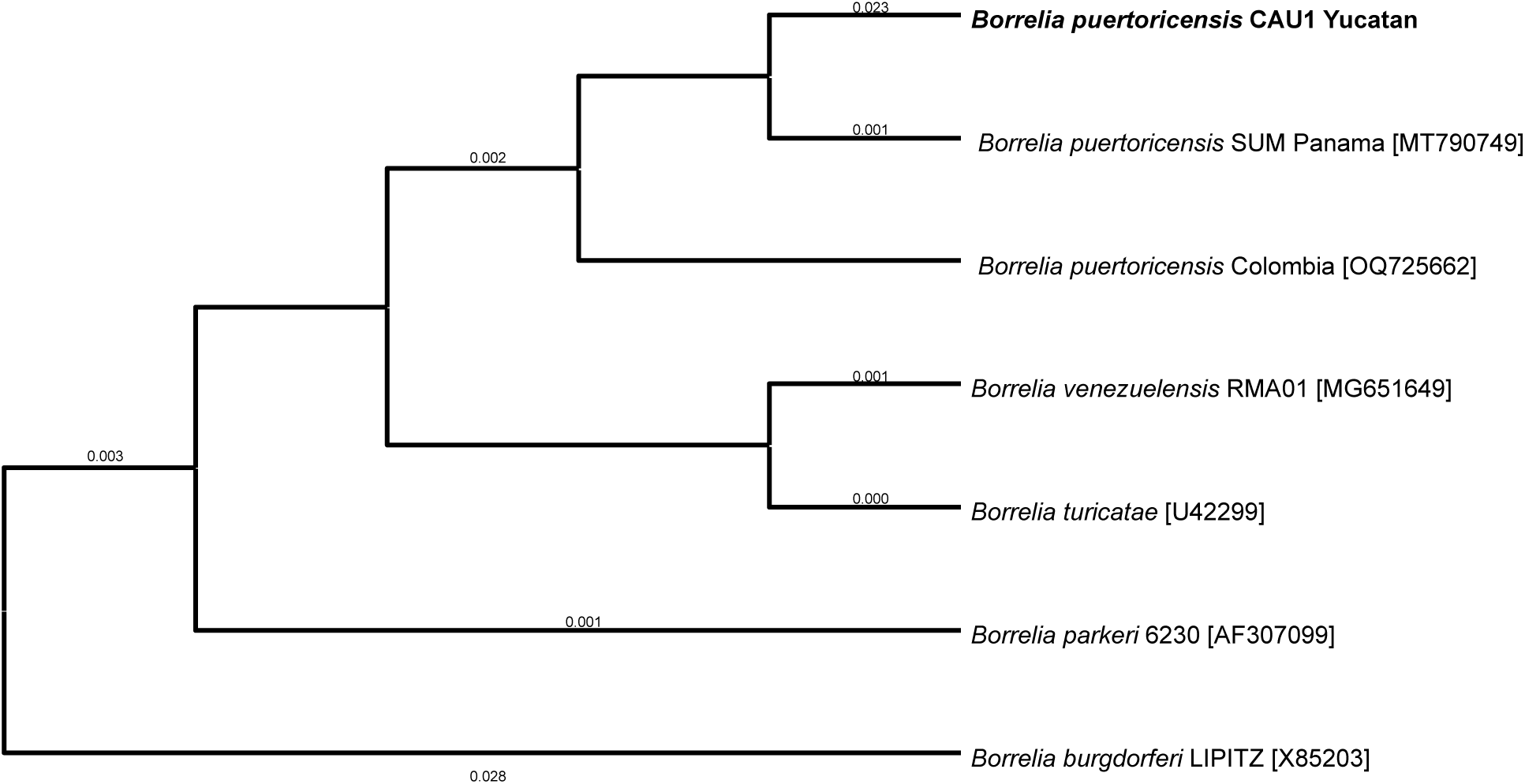
Maximum-likelihood tree constructed from *flaB* partial sequences of various *Borrelia* species. The analysis was performed with the amino acid substitution model Blosum62. Sequences generated in this study are in bold. Numbers represent bootstrap support generated from 1000 replications. The NCBI accession numbers are in brackets and *Borrelia burgdorferi* was used as outgroup.

### Genomic organization of BPUCAU1

Genomic DNA was purified, and spectrophotometry and pulsed field electrophoresis indicated that quality DNA was purified for sequencing [2, 28]. The genome was assembled as reported with minor modifications [27]. The coverages were 439x and 236x, for the short-reads and long-reads, respectively. The assembled and annotated genome was deposited to NCBÍs GenBank (accession numbers CP149102-CP149123). A phylogenomic analysis of 650 single copy genes indicated that the CAU1 isolate was *B. puertoricensis* (Fig 4A). The genome consists of a 928,012 bp linear chromosome, 16 linear plasmids (10,117 bp to 109,690 bp) and 5 circular plasmids (28,746 bp to 59,605 bp) (Fig 4B). Taken together, these results showed that a pool of the argasid ticks collected in Ciudad Caucel are colonized by *B. puertoricensis*.

**Fig 4.**
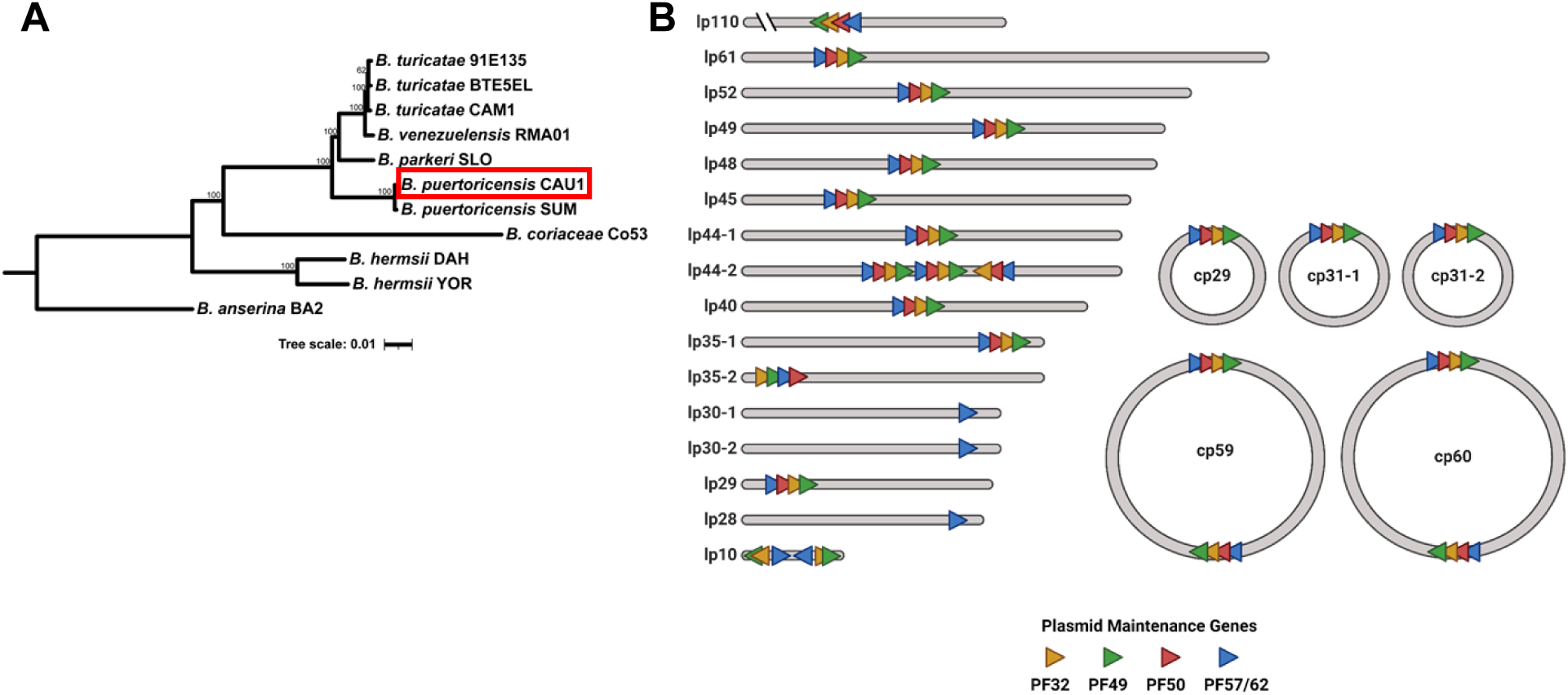
Phylogenomic analysis of *B. puertoricensis* CAU1. The isolate is shown boxed with a red rectangle and additional *Borrelia* genomes were included (A). The tree was generated with an edge-linked proportional partition model with 1,000 ultra-fast bootstraps. Scale bar indicates 0.02 substitutions per site. The genome of CAU1 is shown in (B) and is composed of 16 linear plasmids (lp) and 5 circular plasmids (cp). The numbers indicate the approximate size in base pairs and the colored triangles show the plasmid maintenance genes. PF, plasmid family.

### Identification and analysis of the diagnostic antigen, *Borrelia* immunogenic protein A (BipA)

Since evidence suggests that BipA is a species-specific antigen that can be used to determine vertebrate exposure to RF spirochetes [11], we assessed the CAU1 genome and identified a homolog. An *in silico* translation of *bipA* indicated that it had 97.8% and 71.6% amino acid identity with *B. puertoricensis* SUM (Panama) and *B. turicatae* CAM1 (Mexico), respectively (Table 2) [2, 28]. The detection of a BipA homolog that is divergent from other known species (*B. turicatae*) in Mexico suggests that the antigen could be used serosurviellance studies.

**Table 2.**
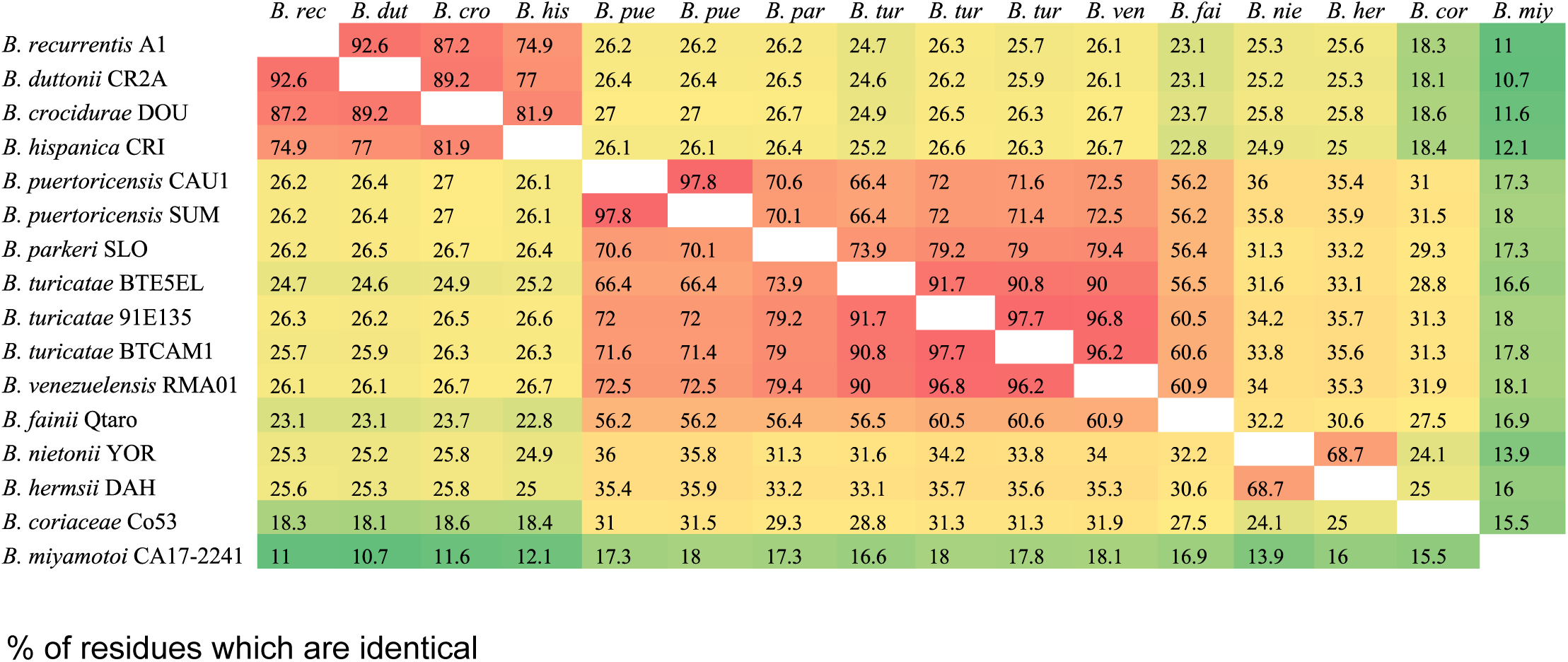
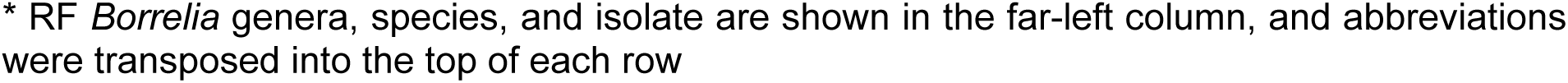
Amino acid identity between BipA homologs.

## Discussion

Despite that twenty-five species of *Ornithodoros* have been reported in Mexico [15], studies of RF spirochetes and their vectors have been neglected and forgotten. Three argasid species, *Ornithodoros turicata*, *Ornithodoros talaje*, and *Ornithodoros dugesi* are known vectors of TBRF spirochetes and transmit *Borrelia turicatae, B. mazzottii* and *B. dugesii,* respectively [3]. Although the presence of *A. puertoricensis* has already been described in Mexico [29] and more recently specimens had been collected in the Mexican state of Veracruz [10], the presence of *Borrelia puertoricensis* has not been demonstrated. In Panama, this spirochete was isolated by using a MyD88(-/-) immunodeficient transgenic murine model, confirming the existence of another *Borrelia* sp. of the RF group and its vector [2].

Isolation of potential pathogens is essential for confirming their circulation in an environment. For RF spirochetes, this is problematic for many reasons including: 1) the vectors ecology; 2) vector misidentification; 3) insufficient diagnostic tests; and 4) difficulty for culturing these bacteria. Recent improvements in culture medium formulations has facilitated the increased success of RF spirochetes [2, 24]. However, the lack of accessible animal models for RF spirochete isolation continued to be a challenge. This is particularly true in laboratories where a genetically immunosuppressed mice are complicated to maintain, such as many Latin America and other resource limited countries.

Burgdorfer and Mavros showed that not all rodent species are competent hosts for RF spirochetes [29], and it was not surprising that immunosuppressed mice were needed for the isolation of *B. puertoricensis*. This *Borrelia* species has been challenging to propagate in Institute of Cancer Research (ICR), BALB/c, C57BL/6, and DBA/2J mouse strains [2]. A study using *A. puertoricensis* collected in Panama initially fed the ticks on ICR mice and spirochetes were undetectable in the blood, but the animal weakly seroconverted to RF *Borrelia* protein lysates [2]. This suggested that the ticks were infected, and they were subsequently fed on a MyD88 -/- mouse. Spirochetes grew to high densities in this mouse strain, which enabled successful spirochete isolation in BSK-IIB medium. While MyD88 -/- mice were not available for our current study, chemically induced immunosuppression was a cost-effective approach. We are currently generating a colony of DBA/2J at the Escuela Nacional de Ciencias Biológicas that can be immune suppressed for isolation of *B. puertoricensis* from the remaining tick cohorts. Additionally, future work will focus on characterizing animal models to study the pathogenesis of *B. puertoricensis*.

There is increasing evidence that *B. puertoricensis* is more prevalent in Latin America than previously thought. This species was identified in argasid tick populations in Panama [2], and DNA from this bacterium was detected in opossum blood in Colombia [30]. Here we have shown that *B. puertoricensis* also circulates in the Yucatan peninsula of Mexico. Our work also demonstrated an extended range of the tick vector, *A. puertoricensis*. More work is needed to understand the public health impact of *B. puertoricensis* on humans, domestic, and wild fauna. In addition to expanded tick collections, we recommend increasing surveillance efforts to determine exposure to RF spirochetes. There have been advances using recombinant BipA (rBipA) as an antigen that can differentiate RF *Borrelia* species causing infection [11]. Future work will generate rBipA from known species circulating in Mexico to aid in defining the public health impact of this neglected pathogen.

## Data Availability

All relevant data are within the manuscript and its Supporting Information files.

## Acknowledgments

We deeply thank Dr. Lilia Dominguez-López and Dr. José Luis González-Quiroz for providing the mice for this study. This work was supported by funds to JAI from Secretaría de Investigación y Posgrado-IPN (20230850, 20240122, 20241496) and by funds provided to JEL from the National School of Tropical Medicine at Baylor College of Medicine and by funds.

